# Effects of hypertension, diabetes and coronary heart disease on COVID-19 diseases severity: a systematic review and meta-analysis

**DOI:** 10.1101/2020.03.25.20043133

**Authors:** Yingyu Chen, Xiao Gong, Lexun Wang, Jiao Guo

## Abstract

**Background:** COVID-19 patients with chronic diseases such as hypertension, diabetes and coronary heart diseases is more likely to worsen, but with mixed results for COVID-19 severity. This meta-analysis is to analyze the correlation between hypertension, diabetes, coronary heart disease and COVID-19 disease severity.

**Methods:** Available data from PubMed, Web of Science, China National Knowledge Infrastructure Database, WanFang Database and VIP Database, were analyzed using a fixed effects model meta-analysis to derive overall odds ratios (*OR*) with 95% *CIs*. Funnel plots and Begg’s were used to assess publication bias.

**Findings:** Of 182 articles found following our initial search, we assessed 34 full-text articles, of which 9 articles with 1936 COVID-19 patients met all selection criteria for our meta-analysis. No significant heterogeneity between studies. There were significant correlations between COVID-19 severity and hypertension [*OR*=2.3 [95% *CI* (1.76, 3.00), *P*<0.01], diabetes [*OR*=2.67, 95% *CI* (1.91, 3.74), *P*<0.01], coronary heart disease [*OR*=2.85 [95% *CI* (1.68, 4.84), *P*<0.01]. Most of the studies in the funnel plot are on the upper part and few on the base part, and are roughly symmetrical left and right. Begg’s test: hypertension (*Z*=-0.1, *P*=1.0), diabetes (*Z*=0.73, *P*=0.466), coronary heart disease (*Z*=0.38, *P*=0.707), all found no publication bias.

**Interpretation:** Hypertension, diabetes, and coronary heart disease can affect the severity of COVID-19. It may be related to the imbalance of angiotensin-converting enzyme 2 (ACE2) and the cytokine storm induced by Glucolipid metabolic disorders (GLMD).

**Funding:** National Natural Science Foundation of China (No. 81830113, 81530102); Major basic and applied basic research projects of Guangdong Province of China (No. 2019B030302005); National key R & D plan “Research on modernization of traditional Chinese medicine” (No. 2018YFC1704200) and Natural Science Foundation of Guangdong Province (No. 2018A030313391)

## Introduction

In December 2019, a cluster of patients with pneumonia of unknown cause was linked to a seafood wholesale market in Wuhan, China.^1^ The pathogen has currently been named severe acute respiratory syndrome coronavirus 2 (SARS-CoV-2).^2^ COVID-19 caused severe acute respiratory syndrome and was associated with ICU admission and high mortality.^3^ The major comorbidities of the fatality cases include hypertension, diabetes, coronary heart disease, cerebral infarction, and chronic bronchitis.^2^

Some research results show that COVID-19 severe patients had a higher proportion of hypertension, diabetes and coronary heart disease. COVID-19 patients combined with cardiovascular disease are associated with a higher risk of mortality.^4^ In this study, we aimed to analyze the correlation between hypertension, diabetes, coronary heart disease and COVID-19 disease severity. This has certain clinical guiding significance for the development of a reasonable comprehensive treatment plan for patients with COVID-19. We strictly followed the inclusion criteria and increased the exclusion criteria to reduce selection bias.

## Methods

### Search strategy and selection criteria

Data sources and searches the PubMed (1970-2020.03.06, English), Web of science (1950-2020.03.06, English), China National Knowledge Infrastructure Database (CNKI, 1979-2020.03.06, Chinese), WanFang Database (1970-2020.03.06, Chinese), and VIP Database (1970-2020.03.06, Chinese) were searched for observational studies of COVID-19. The searches were limited to the English and Chinese languages. Separate search strategies were designed for each database. Search strategies included MeSH, Emtree, Meth terms and free-text, such as “COVID-19”, “SARS Cov-2”, “clinical characteristics”, “clinical features”, and “epidemiological characteristics”. The strategy is as follows:

#1 ‘COVID-19’

#2 ‘SARS Cov-2’

#3 #1 OR #2

#4 ‘clinical characteristics’

#5 ‘clinical features’

#6 ‘epidemiological’

#7 #4 OR #5 OR #6

#8 #3 AND #7

The included studies were selected based on STROBE Statement: describe the setting, locations, and relevant dates, give characteristics of study participants (e.g. the eligibility criteria, diagnostic criteria, the sources and methods of selection), clearly define all outcomes and exposures, etc. The inclusion criteria are listed below. (a) Subjects with laboratory-confirmed COVID-19. (b) It defined the degree of severity of COVID-19 (severe vs. non-severe; or intensive care unit (ICU) vs. non-ICU; or progression and improvement/stabilization) at the time of admission using the American Thoracic Society guidelines for community-acquired pneumonia or Novel coronavirus pneumonia diagnosis and treatment plan (Published by the National Health Committee of the People’s Republic of China, Trial version 4-6). (c) The outcomes have the number of COVID-19 with hypertension, diabetes and coronary heart disease in severe patients and non-severe patients. Literature exclusion criteria: (a) Literatures with review, review, meta-analysis, monographs, conference abstracts, and duplicate literature data. (b) The observation group and the control group did not distinguish the literature of disease severity. (c) Reference quality evaluation scale: Newcastle-Ottawa Scale (NOS) standard, low-quality literature with a score of <6. Two independent reviewers determined whether the observational studies met the eligibility criteria. The reviewers screened the qualified studies in the order of title, abstract and full text. All publications were managed using EndNote X9 software (EndNoteX9), and trials that were excluded in each step were recorded and the reasons were indicated. The data were extracted and collected in specialized spreadsheets. The extracted contents included the number of hypertension, diabetes and coronary heart disease in COVID-19 patients according to disease severity. Discrepancies between reviewers were resolved by a third reviewer.

### Data analysis

Counts and proportions for categorical endpoints were extracted. All statistical analyses were conducted with RevMan 5.3 software. The results for dichotomous outcomes were reported as odds ratios (*OR*) and 95% *CIs*. The Mantel Haenszel (statistical method) and fixed effect models (analysis model) were used to assess outcomes. The I^2^ value was used to evaluate heterogeneity. When I^2^≤50%, the results are considered to be statistically homogeneous, and used a fixed effect model is used for analysis. If the heterogeneity between the studies is large (I^2^>50%), the random effects model is used for analysis. When the research data cannot be combined for analysis or a small probability event occurs, the general description is used for qualitative evaluation or only descriptive analysis is performed. The test level of the meta-analysis was set to α=0.05. Forest plots visually showed effect estimates of the included studies. Funnel plots and Begg’s were used to assess publication bias.

## Results

The process used to select the observational studies followed the Preferred Reporting Items for Systematic Reviews and Meta Analyses (PRISMA) flow diagram. 182 studies were retrieved, and 9 studies were finally included after selection. The detailed screening process is illustrated in Figure 1. All of the studies were conducted in China and published in the Chinese or English language. Baseline data are shown in Table 1. Except for Peng et al ^4^, Which scored 5 points, other studies scored 6 points and above.

**Table 1:** Summary of included studies

| Study | NOS scores | Age, years<br>(mean±SD) |  | Sex (Women/Men) |  | Total |  | Diabetes |  | Hypertension |  | Coronary heart disease |  |
| --- | --- | --- | --- | --- | --- | --- | --- | --- | --- | --- | --- | --- | --- |
|  |  | Severe | Non-severe | Severe | Non-severe | Severe | Non-severe | Severe | Non-severe | Severe | Non-severe | Severe | Non-severe |
| Fang X et al (2020)<sup>5</sup> | 7 | 56.7±14.4 | 39.9±14.9 | 6/18 | 28/27 | 24 | 55 | 4 | 4 | 11 | 5 | 2 | 1 |
| Guan W et al (2020)<sup>3</sup> | 7 | 52.0(40.0-65.0) | 45.0(34.0-57.0) | 73/100 | 386/537 | 173 | 926 | 28 | 53 | 41 | 124 | 10 | 17 |
| Huang C et al (2020)<sup>6</sup> | 6 | 49.0(41.0-61.0) | 49.0(41.0-57.5) | 2/11 | 9/19 | 13 | 28 | 1 | 7 | 2 | 4 | 3 | 3 |
| Liu W et al (2020)<sup>7</sup> | 7 | 66(51-70) | 37(32-41) | 4/7 | 35/32 | 11 | 67 | 2 | 3 | 2 | 6 | NA | NA |
| Wang D et al (2020)<sup>8</sup> | 7 | 66(57-78) | 51(37-62) | 14/22 | 51/61 | 36 | 102 | 8 | 6 | 21 | 22 | 9 | 11 |
| Xiang T et al (2020)<sup>9</sup> | 7 | 53.0±14.0 | 40.6±14.3 | 1/8 | 15/25 | 9 | 40 | 2 | 0 | 4 | 2 | NA | NA |
| Xiong J et al (2020)<sup>10</sup> | 7 | 53.0±16.9 |  | 49/41 |  | 31 | 58 | 6 | 8 | 10 | 16 | NA | NA |
| Yuan J et al (2020)<sup>11</sup> | 7 | 56.4±12.4 | 44.9±16.0 | 13/18 | 105/87 | 31 | 192 | 8 | 10 | 4 | 21 | 0* | 1* |
| Zhang J et al (2020)<sup>12</sup> | 7 | 64(25-87) | 51.5(26-78) | 25/33 | 44/38 | 58 | 82 | 8 | 9 | 22 | 20 | 4 | 3 |

NA=not applicable \*Refers to chronic cardiovascular disease

**Figure 1:**
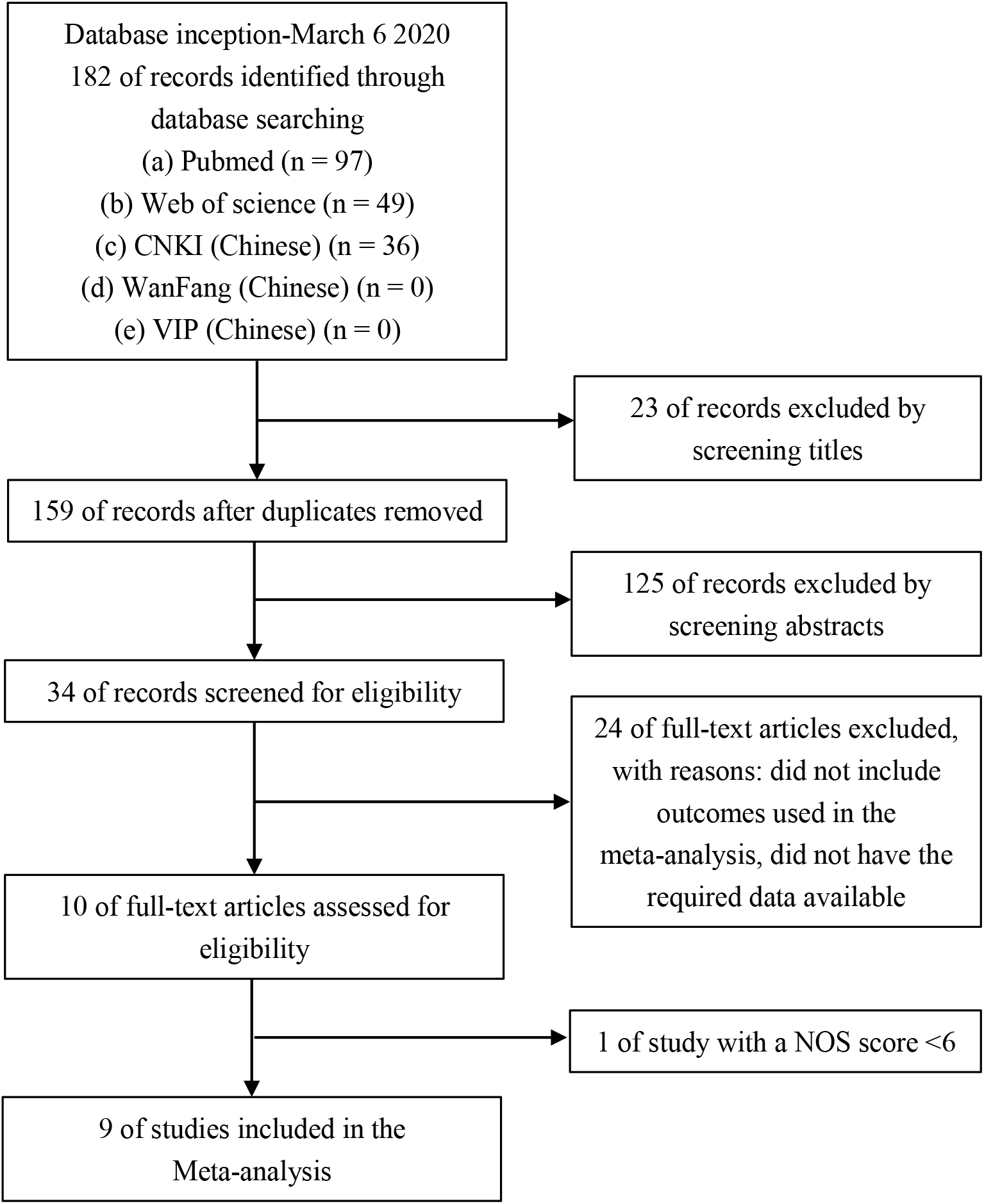
Study selection flowchart

Our study extracted the data of COVID-19 comorbidities included in the literature and listed the distribution of comorbidities. In the included studies, the comorbidities of COVID-19 mainly included hypertension, diabetes, coronary heart disease, chronic liver disease, cancer/malignant tumor, and cerebrovascular disease, chronic obstructive pulmonary disease (COPD). (Figure 2)

**Figure 2:**
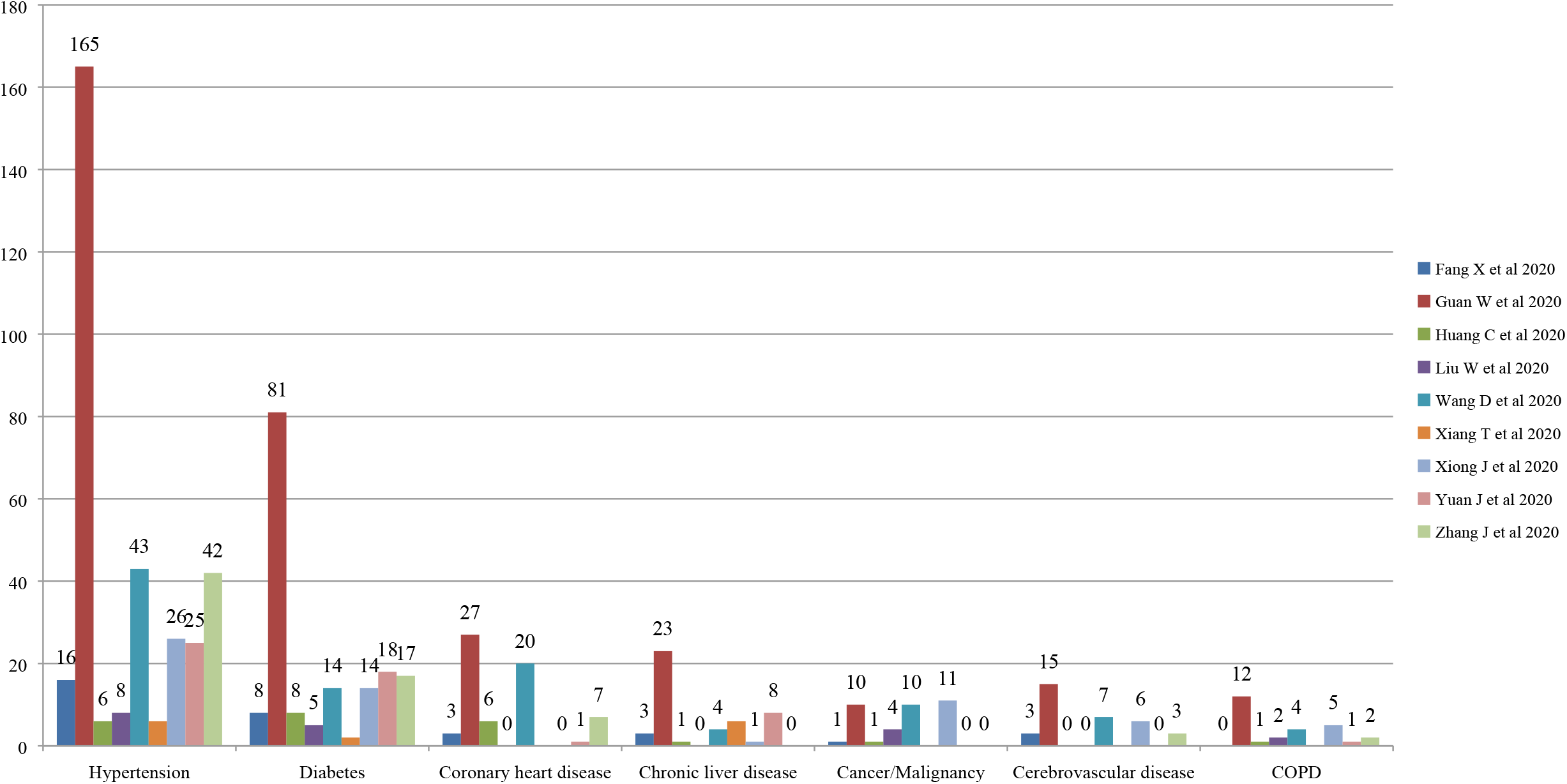
distribution of COVID-19 comorbidities in the included studies

### Efficacy outcomes

Through a systematic review, 9 studies were ultimately included in the statistical analysis (n=1936 participants). Hypertension studies of the heterogeneity (χ2=15.85, *P*=0.04; I^2^=50%), diabetes studies of the heterogeneity (χ^2^=13.78, *P*=0.09; I^2^=42%), coronary heart disease studies of the heterogeneity (χ^2^ =0.61, *P*=0.99; I^2^ =0%). No significant heterogeneity between studies. There were significant correlations between COVID-19 severity and hypertension [*OR*=2.3 [95% *CI* (1.76, 3.00), *P*<0.01], diabetes [*OR*=2.67, 95% *CI* (1.91, 3.74), *P*<0.01], coronary heart disease [*OR*=2.85 [95% *CI* (1.68, 4.84), *P*<0.01]. (Forest plot, Figure 3)

**Figure 3:**
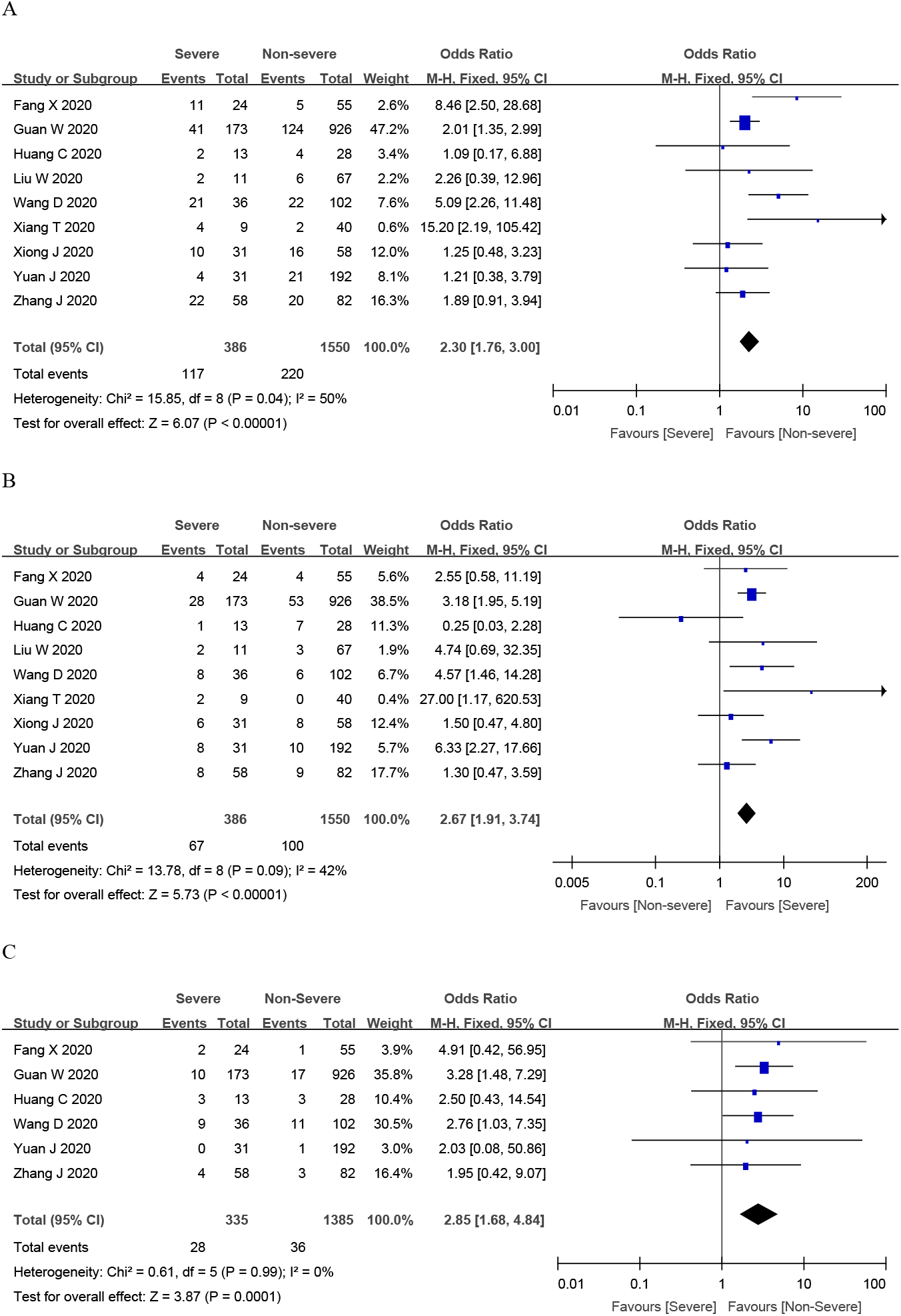
Meta-analysis of the correlations between hypertension (A), diabetes (B), coronary heart disease (C) and COVID-19 severity The X-axis represents the point estimate of odds ratio and corresponding 95% confidence interval; the Y-axis represents the source of study patients; OR, odds ratio; *Cl*, confidence interval.

Most of the studies in the funnel plot are on the upper part and few on the base part, and are roughly symmetrical left and right. Begg’s test: hypertension (*Z*=-0.1, *P*=1.0), diabetes (*Z*=0.73, *P*=0.466), coronary heart disease (*Z*=0.38, *P*=0.707), all found no publication bias. (Funnel plot, Figure 4)

**Figure 4:**
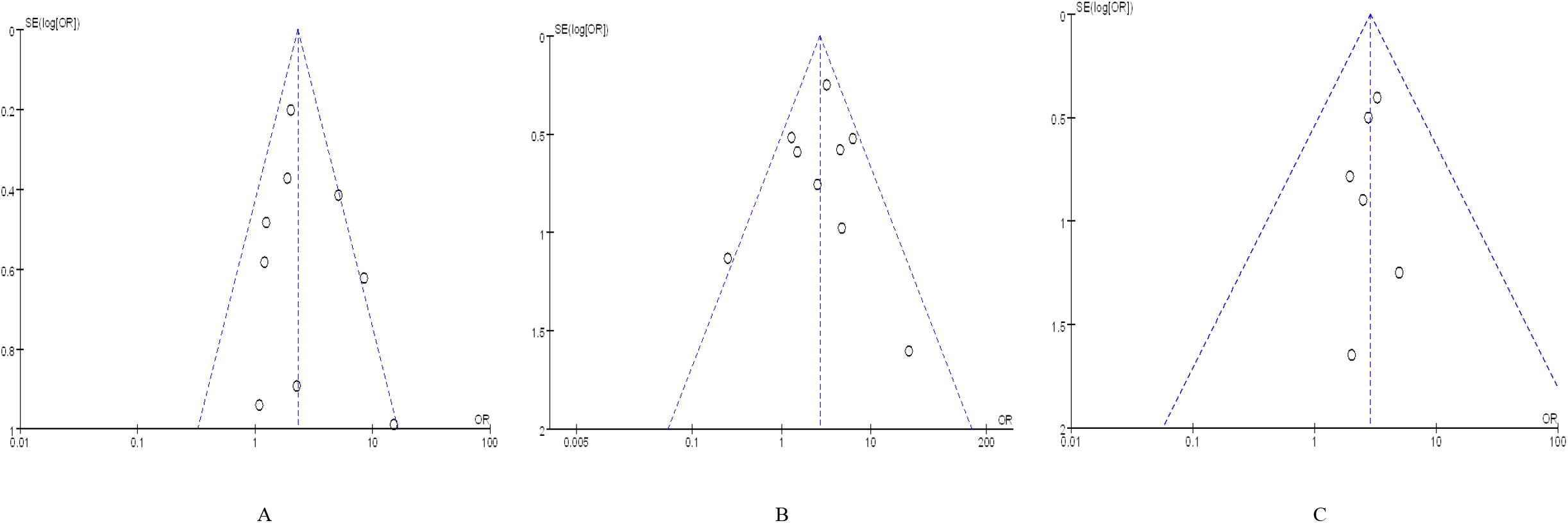
Funnel plot of hypertension (A), diabetes (B), coronary heart disease (C) The X-axis represents the point estimate of odds ratio; the Y-axis represents the standard error (SE); OR, odds ratio; SE, standard error. Begg’s test: hypertension (Z=-0.1, P=1.0), diabetes (Z=0.73, P =0.466), coronary heart disease (Z=0.38, P=0.707), all found no publication bias.

## Discussion

Hypertension, diabetes, and coronary heart disease can affect the severity of COVID-19. It may be related to the imbalance of angiotensin-converting enzyme 2 (ACE2) and the cytokine storm induced by Glucolipid metabolic disorders (GLMD).

### ACE2 imbalance affects COVID-19 severity

Angiotensin-converting enzyme 2 (ACE2), a homologue of carboxypeptidase ACE, is the main active peptide of renin-angiotensin-aldosterone system (RAAS).^13^ By targeting angiotensin II (Ang II), ACE2 has shown protective effects in the cardiovascular system and many other organs^14^, and even protects from severe acute lung failure which infections with severe acute respiratory syndrome (SARS) coronavirus.^15^ Decreased ACE2 shifts the balance in the RAAS to the angiotensin II (Ang II)/AT1R axis, resulting in heart failure progression.^16^ SARS-CoV-2 through spike (S) glycoprotein binds human ACE2 with high affinity.^17^ COVID-19 infection leads to ACE2 more reductions, causing RAAS imbalance and aggravate cardiovascular disease and COVID-19.

In addition, some views consider patients with cardiac diseases, hypertension, or diabetes, who are treated with ACE2-increasing drugs, are at higher risk for severe COVID-19 infection.^18^ However, some reports find ACE inhibitor (ACEI) may have beneficial effects for patients with or at risk for pneumonia, use of lipophilic, but not hydrophilic, ACEI was associated with decreased mortality for patients with community-acquired pneumonia.^19^ Therefore, COVID-19 patients with hypertension, diabetes, or coronary heart disease should be monitored for ACE2-modulating medications, describe the specific medications received, for research to determine the mechanism(s) of effect on COVID-19 diseases severity.

### Glucolipid Metabolic Disorders induce cytokine storms

Hyperlipidemia, diabetes, nonalcoholic fatty liver, atherosclerotic cardiovascular disease and many other metabolic disorders are often combined and affect each other. Professor Jiao Guo from our team proposed the new concept of Glucolipid Metabolic Disorders (GLMD).^20^ GLMD is associated with neuroendocrine disorders, insulin resistance, oxidative stress, chronic inflammatory responses, and intestinal flora disorders.^21-23^ In COVID-19 patient, over activation of T cells lead to the severe immune injury.^24^ Severe COVID-19 patients had higher concentrations of pro-inflammatory cytokines than non-severe patients, suggesting that the cytokine storm was associated with disease severity.^6^ We hypothesized that chronic inflammation in GLMD patients seems to be more activated than in other patients after COVID-19 infection, cause cytokine storm, with hyperinduction of proinflammatory cytokine production^25^, or the cytokine storm caused by COVID-19 may lead to exacerbation of GLMD.

We suggest that a comprehensive analysis of GLMD patients with COVID-19. That is not limited to assess hypertension, diabetes, and coronary heart disease, but also obesity, dyslipidemia, atherosclerosis, and other metabolic disorders etc. Multi-target therapy will help regulate dysfunction of neuronal-endocrine-immune system, which will improve the severity of COVID-19 disease.

### Limitations

It should be acknowledged that the limitations of this meta-analysis may exist in the following points: First, we have included a total of 9 studies, a total of 386 severe patients, 1550 non-severe patients, which is limited to improve the accuracy of the results, only large samples and high-quality research can truly and accurately reflect the correlation between hypertension, diabetes, coronary heart disease and the severity of COVID-19. Second, due to the relatively small number of eligible studies, it is not possible to perform subgroup analysis on the type of severity. In addition, most of the literature does not carry out correlation studies on disease severity time and outcome, so corresponding subgroup analysis cannot be performed. This may also affect its accuracy. Third, there are many factors that affect the severity of COVID-19 disease. In addition hypertension, diabetes and coronary heart disease, it is also related to various factors such as age, other diseases, living environment, economic and health conditions, smoking, viral virulence, and compliance. Most of the included literature is unable to control all factors, which may also affect our results. Finally, as with all meta-analysis, most gray literature is rarely published, and the impact of publication bias on the results of this study cannot be completely ruled out.

## Data Availability

All data included in this study are available upon request by contact with the corresponding author.

## Contributors

YYC and JG designed the study. YYC and XG undertook review activities including searches, study selection, data extraction and calculation, and quality assessment. YYC and XG did the meta-analyses. YYC wrote the manuscript with contributions from JG, LXW and XG. All authors reviewed the study.

## Declaration of interests

We declare no competing interests.

## Acknowledgments

This work was supported by the National Natural Science Foundation of China (No. 81830113, 81530102); Major basic and applied basic research projects of Guangdong Province of China (No. 2019B030302005); National key R & D plan “Research on modernization of traditional Chinese medicine” (No. 2018YFC1704200) and Natural Science Foundation of Guangdong Province (No. 2018A030313391). We are grateful to Yinghua Jin for suggesting thesis writing.

## Notes

### Competing Interest Statement

The authors have declared no competing interest.

### Clinical Trial

NA

